# Immune-checkpoint engagement as predictive biomarkers in clear cell renal cell carcinoma and melanoma

**DOI:** 10.1101/2020.01.15.20017517

**Authors:** Lissete Sánchez-Magraner, James Miles, Claire Baker, Christopher J Applebee, Dae-Jin Lee, Somaia Elsheikh, Shaimaa Lashin, Katriona Withers, Andrew Watts, Richard Parry, Christine Edmead, Jose Ignacio Lopez, Raj Mehta, Stephen G Ward, Peter J. Parker, Banafshé Larijani

**Author notes:** Corresponding Authors: Professors Peter J Parker & Banafshé Larijani Emails. Authors with equal contribution (Lissete Sánchez-Magraner, James Miles). Disclosure: BL and PJP are cofounders of FASTBASE Solutions SL.

## Abstract

Many cancers are termed immuno-evasive due to expression of immuno-modulatory ligands. Programmed death ligand-1 (PD-L1) and cluster of differentiation 80/86 (CD80/86) interact with their receptors, programmed death receptor-1 (PD-1) and cytotoxic T-lymphocyte associated protein-4 (CTLA-4), on tumour infiltrating leukocytes, thus eliciting immunosuppression. Immunotherapies aimed at blocking these interactions are revolutionising cancer treatments, albeit in an inadequately described patient subset.

Our prognostic assay, utilising amplified two-site time-resolved Förster resonance energy transfer (iFRET), quantifies PD-1/ PD-L1 and CTLA-4/ CD80 cell-cell interactions in single cell assays and tumour biopsies. iFRET efficiencies demonstrate, in cell-cell engagement models, that receptor-ligand interactions are significantly lower with anti-PD-1 or anti-CTLA-4 blocking antibodies. In patient samples, iFRET detects immune-cell/tumour-cell interaction variance in different cancers. These results revealed inter-cancer, inter-patient and intra-tumoural heterogeneity of engaged immune-checkpoints, contradicting their ligand expression patterns. Exploiting spatio-temporal interactions of immune-checkpoint proteins defined biomarker functionality for determining whether checkpoint inhibitors are appropriate treatments.

**Statement of Significance:** Quantitative photophysics exploitation in determining immune-checkpoint engagement, as predictive biomarkers in cancers led to revealing inter-cancer, inter-patient and intra-tumoural heterogeneity of the engaged immune-checkpoints. This receptor-ligand interaction did not reflect simple expression patterns of these immuno-modulatory proteins. Our findings may affect immunotherapies aimed at blocking these intercellular interactions in patients.

## Introduction

Disproportionate immune-system activation can result in profound pathologies and there are, therefore, regulatory mechanisms in place to maintain homeostasis [1]. Interactions referred to as immune-checkpoints are critical in this, avoiding immune-cell related collateral damage in pathogenic responses and in suppressing autoimmunity. Inhibitory receptors presented by immune cells, T-cells in particular, include programmed death receptor-1 (PD-1) and cytotoxic T lymphocyte antigen-4 (CTLA-4) [2]. Cancers exploit these physiological mechanisms to avoid immune-attack by expressing inhibitory receptor ligands, programmed death-ligand 1 (PD-L1) and cluster of differentiation 80/86 (CD80/86) [1]. The CTLA-4 receptor is a homolog of the immune-activating CD28 receptor, both of which are found on T-cells and possess CD80 and CD86 as ligand partners. CTLA-4, however, provides a higher affinity binding site for CD80/86 and engagement with CD80/86 inhibits cell proliferation and interleukin-2 (IL-2) secretion by T cells. The PD-1 immune checkpoint, limits later immune responses primarily in peripheral tissue [3].

There are a number of approved, monoclonal antibodies (mAbs) designed to reinstate immune-mediated tumour destruction in immunogenic cancers, by inhibiting these immune-checkpoint interactions [4]. In part through the generation of neo-antigens, immunogenicity is strong in non-small-cell-lung-cancer (NSCLC), renal cell carcinomas (RCCs), melanoma, classical Hodgkin lymphoma, head and neck squamous cell carcinoma and urothelial carcinoma, all of which show varying degrees of response to immune-checkpoint interventions [4]. Notwithstanding some remarkable successes with immune-checkpoint inhibitors, the response rates in patients with these tumour types are estimated to be limited to 15%, with the majority of patients showing no response. There is therefore an unmet medical need to identify biomarkers that distinguish potential responders from non-responders to ensure that non-responders are not exposed to the side effects of the drugs for no therapeutic benefit. The development of different PD-L1 immunohistochemistry (IHC) diagnostics utilising proprietary antibodies has resulted in four FDA-approved and CE-*in vitro* diagnostics (IVD)-marked assays, each linked to a specific drug and scoring system. However, it has become clear that expression of inhibitory ligands, namely PD-L1 is not an accurate diagnostic marker for use in predicting patient prognosis and reaction to treatment. A recent study observed that NSCLC patients demonstrated an increase in response to anti-PD-1 agent, pembrolizumab, in patients eliciting a tumour proportion score (TPS) greater than 50%. Nevertheless, the response reached only 41% [5]. Moreover, a different study assessed the efficacy of PD-1 or PD-L1 inhibitors in different neoplasia (primarily lung cancer but also renal cancer and melanoma) in PD-L1 negative and PD-L1 positive cancers. Critically, benefit was seen in patients within the PD-L1 negative group, clearly exposing the failure of PD-L1 expression to determine which patients should receive immune-checkpoint inhibitors [6].

As immune-cell/tumour-cell interplay via immune-checkpoints is a prominent mechanism for tumour immune-evasion and survival, interaction status may present as a valuable prognostic biomarker, replacing conventional protein expression readouts for stratifying patients to immune-checkpoint interventions. We have thus implemented a two-site, amplified, Förster resonance energy transfer detected by Fluorescence Lifetime Imaging Microscopy (A-FRET/FLIM) assay to enhance specificity and the dynamic range. This provides an in-depth analysis of cell-cell contact and can further define patient cohorts considered for immune-checkpoint inhibitor therapy. The distance criterion (1-10nm) of FRET can be exploited to determine receptor-ligand interaction. Critically, data are unaffected by absorption by the sample, sample thickness and photo bleaching [7]. The methodology utilises tyramide signal amplification (TSA), as described by Toda et al., 1999 [8], to enhance signal-noise ratio. A-FRET has been successful in determining Akt activation status in primary breast carcinoma and clear cell RCC (ccRCC) biopsies, in which IHC intensity ratios failed to do so, and correlated activation with poor patient outcome [9, 10]. In the above studies FRET Efficiency (E_f_) has been utilised as the parameter that precisely determines molecular activation and interaction status. Here, A-FRET was applied to determine intercellular ligand/receptor interactions; this is referred to as immune-FRET (iFRET).

Here we have investigated if iFRET could determine cell-cell interaction in single cell assays involving complimentary receptor and ligand expressing cells. When PD-1/PD-L1 proximity was assessed using the two-site proximity ligase assay (PLA), it was shown that the assay did not determine interaction state. The success of iFRET in quantifying intercellular interaction in cell assays led to its application to formalin fixed paraffin embedded (FFPE) patient tumour biopsies. iFRET detected immune-cell/tumour-cell interaction in a variety of cancer types, but revealed high levels of inter-cancer, inter-patient and intra-tumoural heterogeneity. Furthermore, iFRET was able to determine significant differences in patient outcome, when assessing PD-1/PD-L1 interaction in melanoma, whereas classical expression-based methods did not. These unprecedented results illustrate that iFRET can be used for a more precise measurement of PD-1/PD-L1 in clear cell renal cell carcinoma and melanoma carcinomas.

## Results

### Development and validation of a novel amplified-FRET imaging assay for determining immune-checkpoint interaction in single cell assays

The Promega Blockade Bioassay, originally designed to measure the antibody blockade of PD-1/PD-L1 and CTLA-4/CD80 interaction by luminescence, was adapted for an iFRET protocol with the aim of verifying the technique for detecting intercellular interaction of these ligand/receptor pairs. The intercellular interaction and FRET efficiencies were determined (Figures 1 and 2).

**Figure 1:**
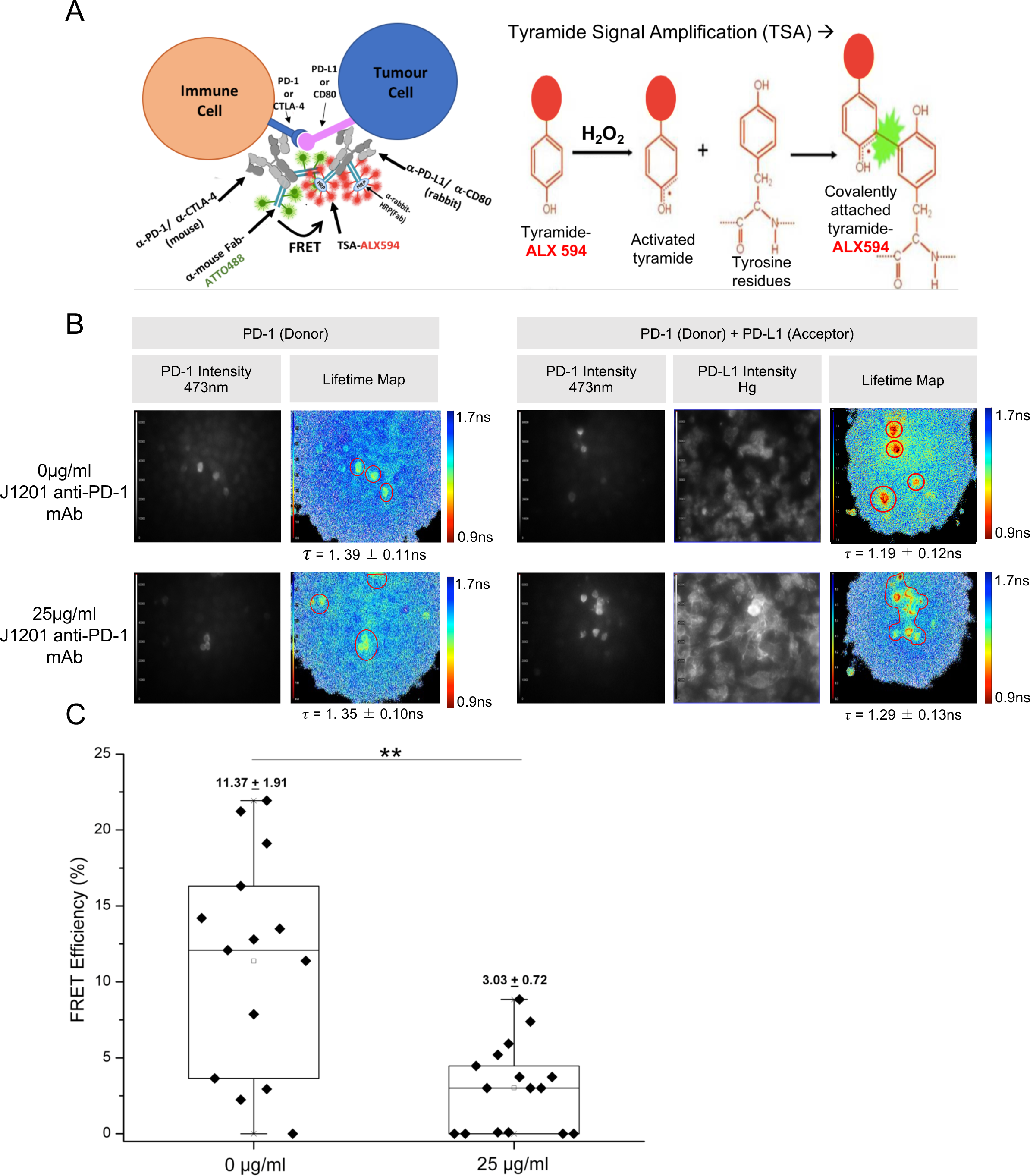
iFRET detects and quantifies PD-1/PD-L1 interaction in single cells. A) Schematic diagram showing the principle of the A-FRET assay. PD-1 and PD-L1 are labelled with primary antibodies. F(ab) fragments are used as secondary antibodies, conjugated to ATTO488 (donor) or HRP/Alexa594 (acceptor). B) Black and white panel images represent PD-1 (ATTO488) and PD-L1 (ALEXA594) intensity and therefore expression. Pseudo-colour lifetime maps indicate the *τD* (PD-1; ATTO488) and *τD*+A (PD-1; ATTO488 + PD-L1; ALEXA594) conditions in the absence (upper) and presence (lower) of anti-PD-1 mAb. Regions of acquisition are outlined in red and localised donor lifetimes (*τ*) are indicated. C) Box and Whisker plot compares FRET efficiency values in the absence and presence of 25μg/ml J1201 anti-PD-1 mAb (added prior to cell fixation). Mean FRET efficiencies ± SEM are indicated. Mann Whitney U analysis determined statistical differences between treated and untreated cells (**, p=0.004).

**Figure 2:**
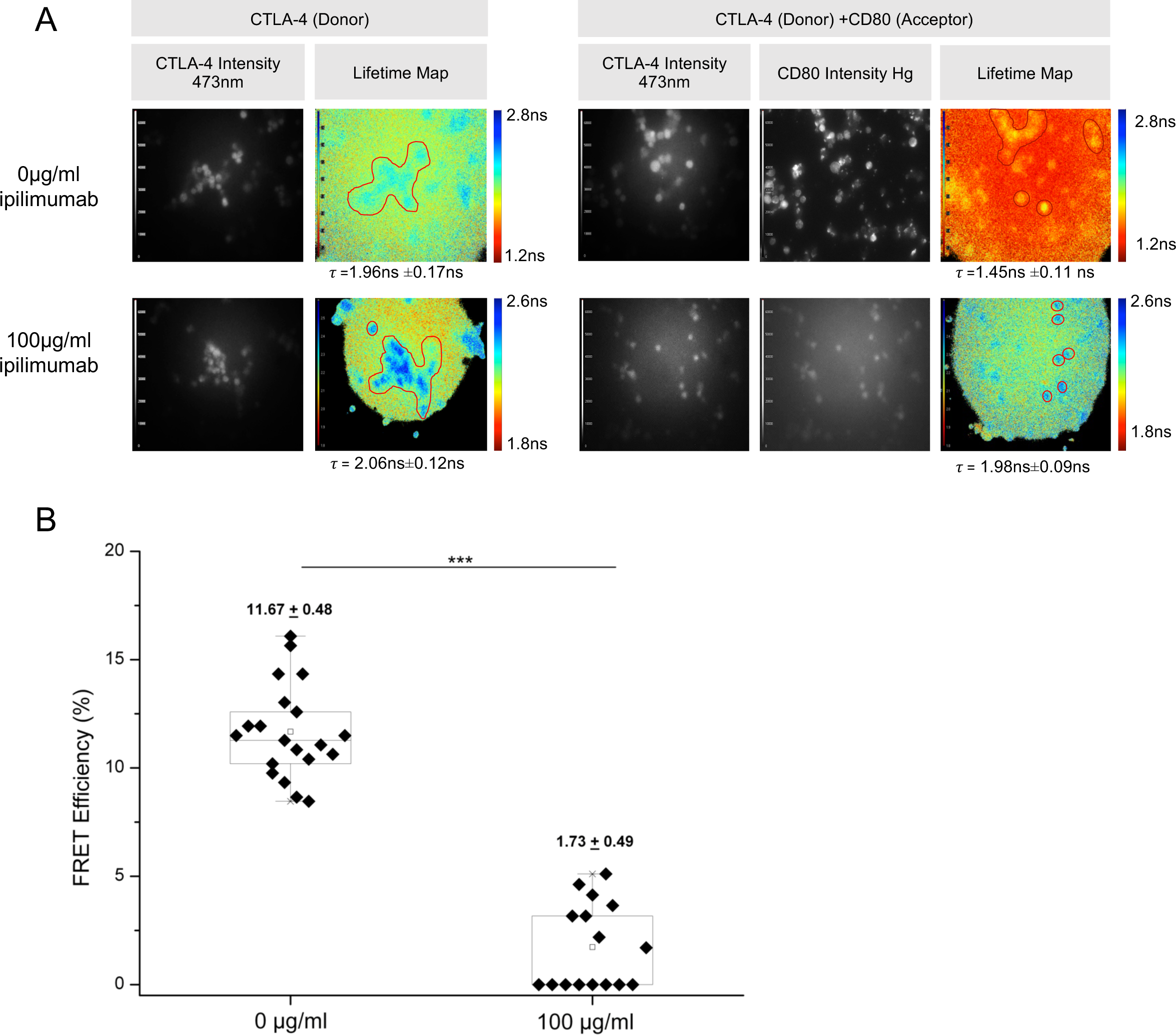
iFRET precisely determines CTLA-4/CD-80 interaction. A, Panels show the intensity images of CTLA-4 (ATTO488) and CD80 (Alexa594) and lifetime maps of *τD* (CTLA-4; ATTO488) and *τD*/A (CTLA-4; ATTO488 + CD80; ALX594) conditions in the absence (upper) and presence (lower) of anti-CTLA-4 drug, ipilimumab (added prior to cell fixation). Regions of acquisition are outlined in red and localised *τ* are indicated. B, Box and Whisker plot compares FRET efficiency values in the absence and presence of 100μg/ml ipilimumab (maximal inhibition). Mann Whitney U analysis determined statistical differences between treated and untreated cells (***, p=3.27 e-7).

Figure 1 illustrates intercellular interaction of PD-1 and PD-L1, on Jurkat and CHO-K1 cells, using iFRET. Cells were not permeabilised and therefore the observable interaction was that of two membrane-bound, extracellular proteins. FRET efficiency values were significantly (**) higher (p=0.004) in the absence of blocking antibody when compared to cells treated with 25μg/ml anti-PD-L1 mAb J1201. The findings suggest that the decrease in donor lifetime is due to the specific interaction of PD-1 and PD-L1.

In order to determine the relationship between PD-1/PD-L1 expression and their interaction, fluorophore intensity was correlated with FRET efficiency. PD-1 intensity did not correlate with FRET efficiency in the presence or absence of blocking antibody, with r_2_ values of 0.22 and 0.58 respectively (Supplementary Figure 1A). PD-L1 expression also did not correlate with FRET efficiency, in the presence or absence of blocking antibody, with r_2_ values of 0.054 and 0.19 respectively (Supplementary Figure 1B).

Intercellular CTLA-4 and CD80 interaction, in Jurkat and Raji cells, was also assessed using iFRET (Figure 2). A substantial decrease in lifetime (ns) was observed between D and D/A conditions in the absence of ipilimumab. In agreement with this, FRET efficiency values were significantly (***) higher (p=3.27×10^−7^) in the 0μg/ml ipilimumab condition compared to 100μg/ml ipilimumab condition suggesting the interaction seen is a specific interaction between CTLA-4 and CD80. As with the PD-1/PD-L1 pair, there was no correlation between CTLA-4/CD80 expression and interaction status. CTLA-4 expression did not correlate with FRET Efficiency in the absence or presence of blocking antibody, with r_2_ values of 0.35 and 0.17 respectively (Supplementary Figure 2A). CD80 expression also failed to correlate with FRET efficiency in the absence or presence of blocking antibody, with r_2_ values of 0.11 and 0.33 respectively (Supplementary Figure 2B).

For these iFRET immune-checkpoint engagement assays, it is evident that they require proximal engagement of the respective ligand receptor pairs and that the extent of this engagement is, as would be anticipated, not a simple correlate of expression levels.

### PLA fails to identify PD-1 and PD-L1 interaction in PD-L1 negative ccRCC sample as determined by iFRET

To benchmark the effectiveness of the iFRET assay we compared this to a Proximity Ligation Assay (PLA), which in principle can also visualise PD-1 and PD-L1 within close proximity. To achieve this comparison analyses were run on, on sequential sections of ccRCC tissue. The samples utilised in both the PLA and iFRET assays were sequential tissue sections. Prior to the investigation, samples were determined PD-L1 positive (>1%) or negative (<1%) using the Roche VENTANA PD-L1 (SP142) assay.

PLA allowed the visualisation of PD-1 and PD-L1 within close proximity (Supplementary Figure 3A). The PD-L1 positive ccRCC sample labelled with anti-PD-1, anti-PD-L1 and PLA +/-probes produced the greatest PLA signal. However, PLA signals were observed across both experimental and control groups (normal renal tissue) possibly due to PLA only determining close proximity as opposed to interaction.

**Figure 3:**
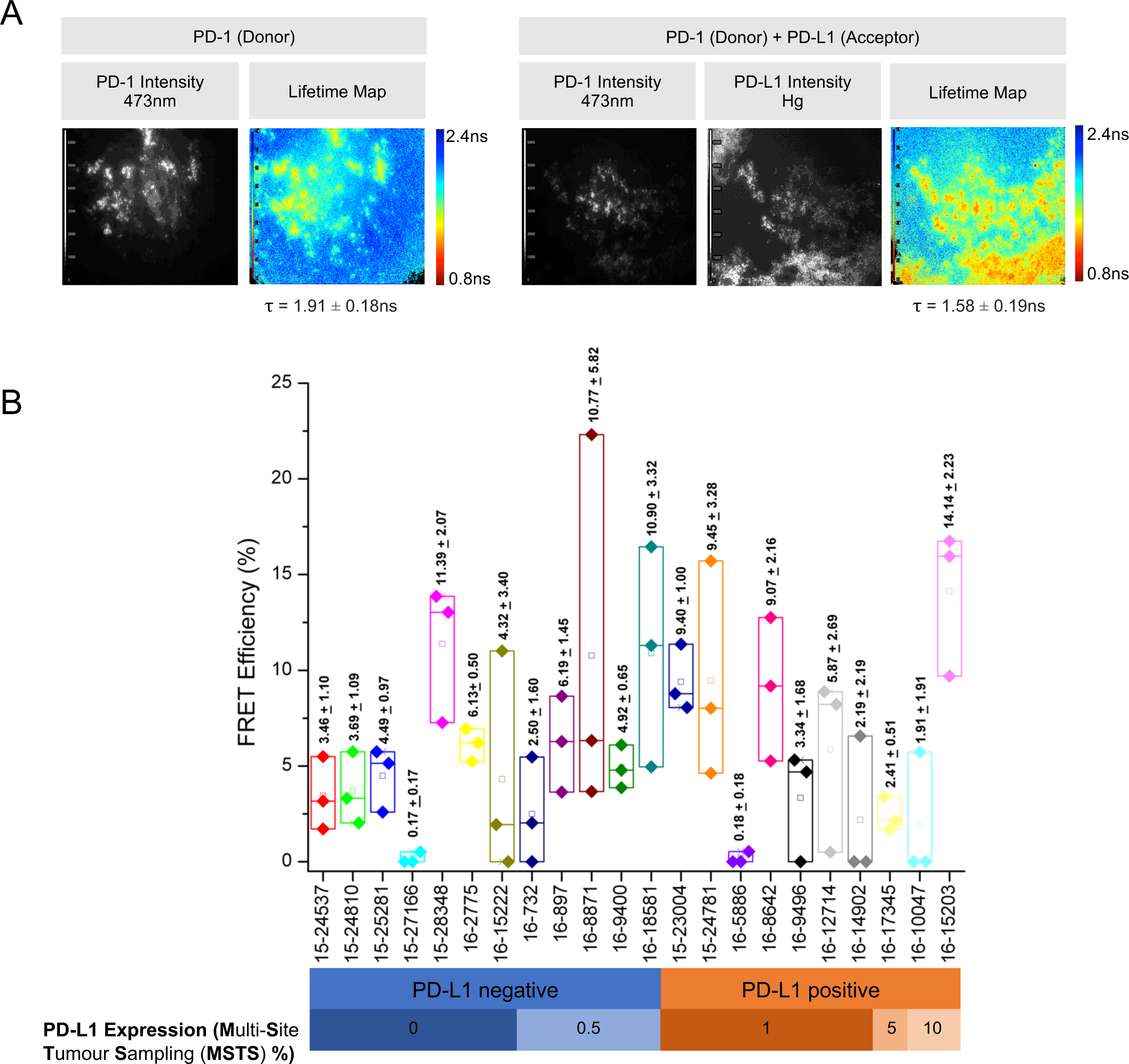
iFRET detects heterogeneity of PD-1 and PD-L1 interaction in FFPE ccRCC. A) Intensity images and lifetime maps (pseudo-colour scale) of FFPE human ccRCC patient sample 16-15203 labelled with D (PD-1; ATTO488) or D+A (PD-1; ATTO488 + PD-L1; Alexa94). Coincidence is observed between PD-1 and PD-L1 situation in the D/A condition. A decrease is observed from *τD* to *τD*A in the example provided. The localised *τ* value ± standard deviation is indicated. B) Box and whisker plots represent the upper quartile of FRET efficiency values for each patient tissue sample. The mean values ± SEM are indicated.

In the sample illustrated, iFRET identified an area of high interaction between PD-1 and PD-L1 (Supplementary Figure 3B). However, when analysing PLA labelling of PD-1 and PD-L1, this PD-L1 negative sample displayed an apparently minimal interaction state. The observations suggest that iFRET provides greater sensitivity, thus allowing the identification of tumour-mediated immune-suppression in patients otherwise considered as PD-L1 negative.

### PD-L1 expression does not correspond to interaction status of PD-1 and PD-L1 in ccRCC

Following iFRET optimisation and benchmarking, we assessed the interaction of PD-1 and PD-L1 in FFPE ccRCC patient samples. The series included samples from 22 patients considered as PD-L1 negative (<1%) or positive (>1%), using the Roche VENTANA PD-L1 (SP142) assay and multi-site tumour sampling (MSTS). The mean FRET efficiency varied from 0.17% to 14.14%, suggesting the iFRET assay is able to quantitatively detect the PD-1 and PD-L1 interaction in patients over a good range. Differing levels of interaction between Tumour Infiltrating Leukocytes (TILs) and tumour cells was observed in samples within those classed as PD-L1 positive or negative (Figure 3). Notably, PD-L1 expression, classified by MSTS, did not correlate with the interaction status of PD-1 and PD-L1 as determined by iFRET.

### iFRET identifies and stratifies patient response to immunotherapy in melanoma

After analysing PD-1/PD-L1 interaction in ccRCC tissue, the interaction status in 176 patients was assessed in melanoma TMAs. The cohort was predominantly male with a split of 102M/71F and a mean age of 66.1 years. 25% of patients had stage one tumours, 43.5 had stage two tumours, 9.4% had stage three tumours and 22.1% had stage four tumours. Tumour infiltrating lymphocytes were absent in 39 patients, 101 patients had focal infiltration with 30 patients experiencing extensive infiltration (Supplementary Table 1).

Figure 4A is the Haematoxylin and Eosin (H&E) staining of a primary cutaneous malignant melanoma; the left-hand panel shows the H&E staining of a patient with high tumour infiltrating lymphocytes (circled), this patient had a FRET efficiency of 26.20%. The right-hand panel shows a patient with minimal tumour infiltrating lymphocytes, this patient had a lower FRET efficiency of 3.50%. Figure 4B shows FLIM images of the sample of patient 390, wherein intensity maps demonstrate the expression of PD-1 and PD-L1. The lifetime maps identify the donor lifetime using a pseudo-colour scale, where blue indicates a higher lifetime (3.5ns) and red indicates a lower lifetime (0.5ns). Despite a high expression of PD-L1 in this patient’s sample, a low change in lifetime was observed; donor lifetime alone was 1.95±0.16ns and slightly decreased to 1.88±0.15ns in the presence of the acceptor, resulting in a low FRET efficiency (Ef%)= 3.50%, meaning that the ligand and receptor were interacting at a low level. Conversely, Figure 4C shows the sample of patient 131. As observed in the sample of patient 390, patient 131s’ sample demonstrated a high level of PD-L1 expression. However, unlike patient 390, patient 131 displayed a high interaction state between ligand and receptor, with the donor lifetime decreasing from 2.22±0.19ns to 1.64±0.15ns when in the presence of the acceptor, with a resulting FRET efficiency of 26.20%. These results infer that PD-L1 expression does not correlate with PD-1/PD-L1 interaction.

**Figure 4:**
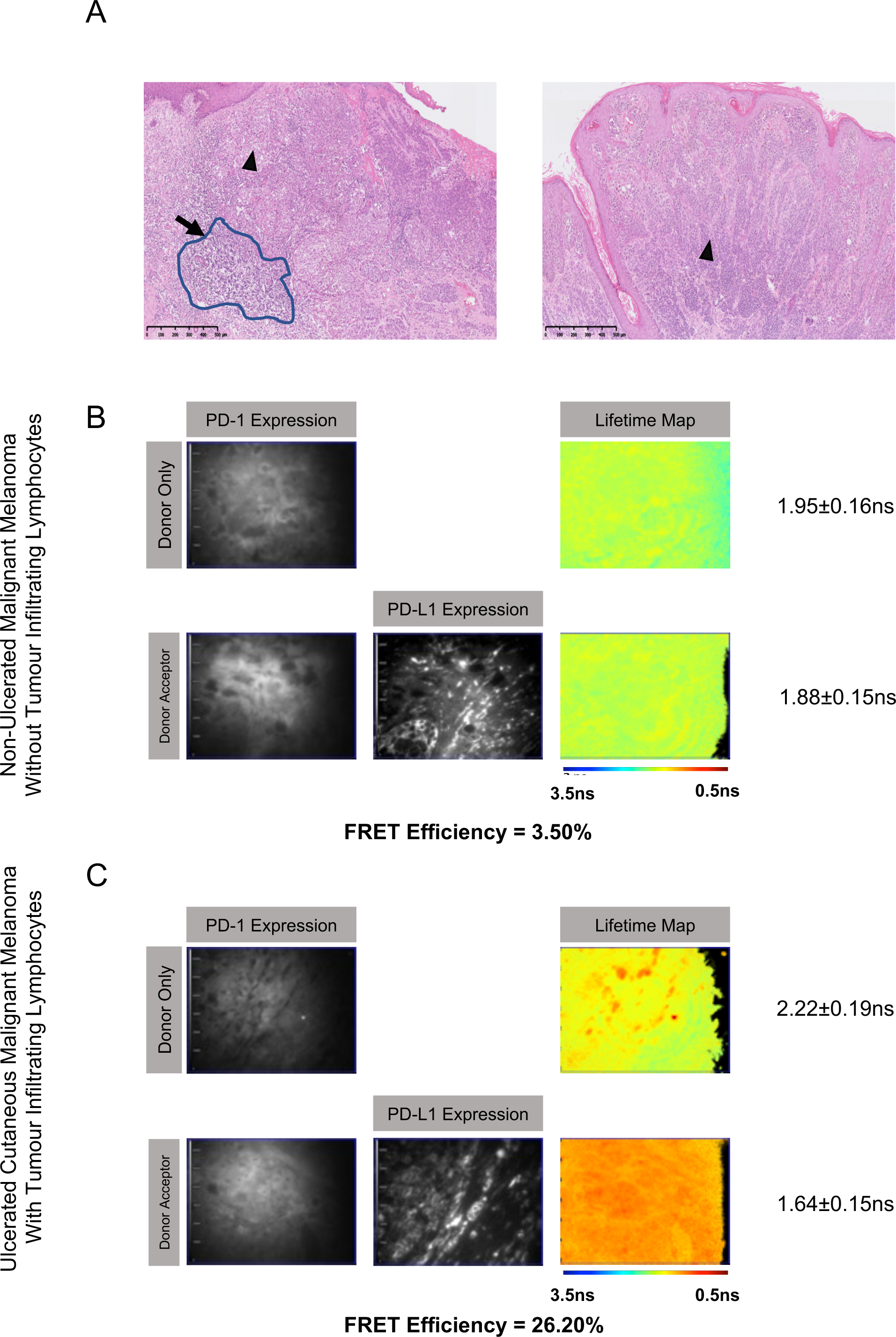
H&E Staining of high- and low-grade tumours does not correlate with PD-1/PD-L1 interaction state. A) The left-hand image shows the H&E. staining of an ulcerated cutaneous malignant melanoma (triangle) with brisk tumour infiltrating lymphocyte (arrow) and high FRET efficiency (26.20%). The area of tumour with lymphocyte infiltration is circled. The right-hand image shows non-ulcerated malignant melanoma with absent tumour infiltrating lymphocytes (triangle) and low FRET efficiency (3.50%), B) Fluorescence lifetime imaging microscopy (FLIM) images show a melanoma with a low PD-1/PD-L1 interaction state. Expression images, based on PD-1 or PD-L1 intensity, show the presence of the receptor and ligand, however, the lifetime map shows no change in pseudo-colour, indicating a lifetime change of 1.95±0.16ns to 1.88±0.15ns and thus a low interaction state. C) FLIM images show a melanoma sample with a high PD-1/PD-L1 interaction state. Again, the expression maps show the presence of PD-1 and PD-L1 as in panel B, however the change in pseudo-colour represents a change in lifetime of 2.22±0.19ns to 1.64±0.15, indicating a high interaction state.

Figure 5A shows the distribution of FRET efficiencies for the 176 melanoma patients. The FRET efficiencies ranged from multiple patients not exhibiting any PD-1/PD-L1 interaction state (FRET efficiency of 0%) up to a maximal interaction state of 31.30% observed in one patient. This demonstrates the high dynamic range of the iFRET assay which is able to detect and quantify inter-tumour heterogeneity amongst a patient cohort.

**Figure 5:**
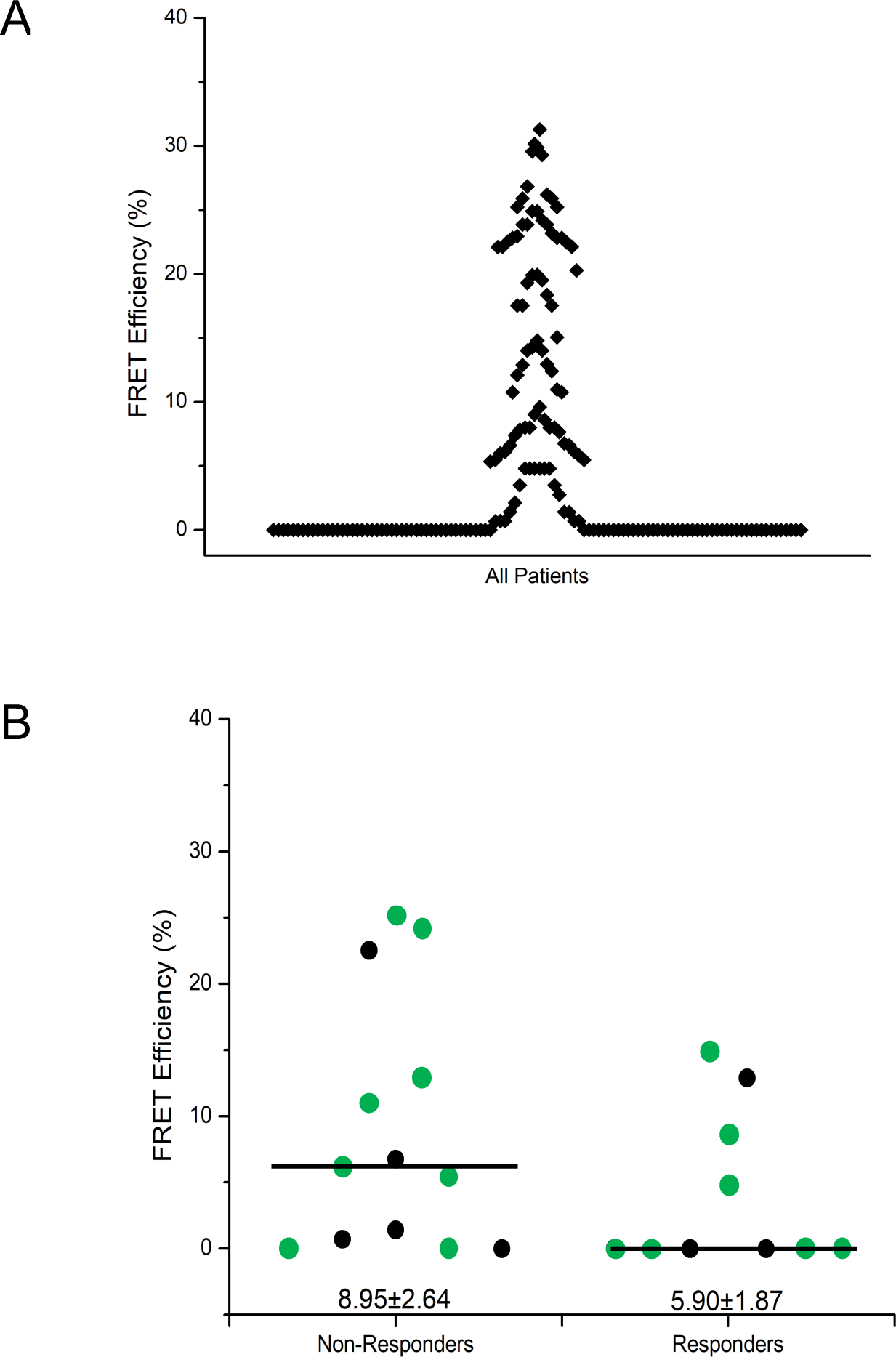
iFRET identifies and stratifies patient response to immunotherapy in melanoma. A) iFRET quantifies inter-patient heterogeneity in melanoma. The high dynamic range of iFRET allows the quantification of FRET efficiencies from 0% to a maximum of 31.30%. B) Mann-Whitney plots correlate patients’ FRET efficiency in 23 immunotherapy-treated patients (bars represent median with the mean ± standard error of the mean indicated underneath each plot). Responders were defined as those who progressed within 10 months with responders not progressing for over 40 months, if at all. Responders exhibit lower FRET efficiencies (PD-1/PD-L1 interaction state) compared to non-responders and higher tumour infiltrating lymphocytes (TILs). The opposite was observed in responders.

We compared the response to clinical intervention of the treated subset of patients. From the 176 patients only 23 high grade melanoma patients were treated with immunotherapy and their FRET efficiencies were correlated with whether they responded or did not respond to therapy. Patient response to immunotherapy was assessed by evaluating time taken from the start of immunotherapy until relapse. Non-responders are categorised as those progressing within 10 months (13); by contrast responders did not relapse until >40 months (2) if at all (8). Patients who responded to therapy had lower FRET efficiencies indicating a lower interaction state. Conversely, those patients who did not respond showed a higher FRET efficiency and higher interaction state (Figure 5B). Furthermore, when the absence or presence of tumour infiltrating lymphocytes (TILs) was considered out of the 23 treated patients, the 12 who responded to immunotherapy treatment had a population of patients with low TILs. The non-responders had in general an enhanced number of TILs and higher FRET efficiency at diagnosis.

## Discussion

This study has demonstrated the successful application of iFRET to detect intercellular ligand-receptor interactions. The method applied combines a two-site time-resolved FRET assay and signal amplification, with a tissue preparation time identical to that of IHC approaches. The high-throughput frequency domain FRET/FLIM imaging platform allowed mapping and automated acquisition of data from both cell cultures and arrayed tissue samples, thereby creating a straightforward procedure for non-specialised personnel. The methodology was exemplified for the interaction status of two immune-checkpoint pairs, PD-1/ PD-L1 and CTLA-4/CD80, in single cell assays (Figures 1 and 2) and biopsy tissue samples from patients with ccRCC (Figure 3) and malignant melanoma (Figure 4).

The validation of the method in single cell assays where manipulation of ligand-receptor interactions can be specifically manipulated, has provided the confidence to assess these complexes in patient biopsies. The additional controls with respect to the use of single antibodies and single secondary reagents add further to the validity of the assay platform and of course are variables than can be applied routinely to patient biopsies. Comparison with the PLA technology provided evidence that the latter did not perform as well in these settings in defining complex formation. By its very design, the iFRET methodology elaborated here provides both a measure of ligand-receptor engagement and the spatial resolution of this engagement. Importantly this is readily achieved in routinely fixed samples from patient biopsies, offering great promise in being able to inform on the more detailed behaviour of these interactions and their distribution within pathological settings. This is well illustrated here with the observed heterogeneity seen not simply between patient biopsies but within individual biopsies. This heterogeneity may reflect tumour heterogeneity and differential patterns of reprogramming of the tumour microenvironment, playing out in modified immune-suppressive ligand presentation and/or variability in the degree of immune-cell infiltration.

A lack of correlation between the extent of PD-1/PD-L1 complex formation and the expression levels of these two proteins was evident in both patient cohorts studied. This has implications for the use of simple expression levels in stratifying patients for treatment. Blockade of interaction would be predicted to be effective in contexts where engagement occurs and is responsible for the immune privileged state of the tumour. Hence engagement would *a priori* be a criterion for treatment. However, this simple relationship is predictably more complicated. While the numbers of immune-checkpoint inhibitor treated patients analysed here is small, the trend observed in these metastatic melanoma patients is that those displaying the highest levels of complex formation, actually show the least effective response to treatment. What extent is this a function of the extent of target engagement in these patients? Critical questions such as this now require a longitudinal study in a larger cohort of patients.

iFRET can be exploited to monitor other intercellular protein interactions and there are ongoing developments designed to capture related immune modulatory interactions pertinent to cancer and emerging cancer treatments. This provides the potential for iFRET to become a useful predictive tool informing on the nature of the tumour immune-privileged state. However, as a principle, it is clear that this approach has capabilities beyond immune-tumour cell interactions and the broader uptake of the approach promises to be informative in many research (e.g. axon guidance) and clinical (e.g. angiopathies) settings.

The exemplification of iFRET in tumour settings opens up opportunities to move beyond the cataloguing of cell phenotypes *in situ* and add functional attributes to our patient data inventory, impacting clinical decisions. This is a routine parameter for small molecule inhibitors targeted at driver mutations and we suggest it should become a routine for these more complex biotherapeutic interventions.

## Material and Methods

### Pathology

Biopsies from renal cell carcinoma patients, diagnosed and treated at the Cruces University Hospital, Bizkaia, Spain, were graded and staged within the study. All patients were informed about the potential use of their resected tumours for research and consented to this. This study was approved by the Ethical and Scientific Committee (CEIC-Euskadi PI2015060). The International Society of Urological Pathology (ISUP) 2013 tumour grading system [11] was used to assign each sample using routine haematoxylin and eosin (H&E) staining. Tumours were graded and grouped as low (G1/2) and high (G3/4) grade for consistency. To assess PD-L1 expression, a multi-site tumour sampling (MSTS) method was used which samples more areas of a tumour with the aim of overcoming the problems of tissue heterogeneity [12]. Samples were determined PD-L1 positive (>1%) or negative (<1%) using the Roche VENTANA PD-L1 (SP142) assay.

Cases of malignant melanoma were selected from all patients diagnosed with malignant melanoma (MM) between June 2003 and February 2017 at Nottingham University Hospital. The main selection criterion was tumours having a Breslow thickness of >1mm. Patients gave informed consent for their specimens to be stored and used for research. Patient clinicopathological data was obtained from Nottingham University Hospital PAS, WinPath and NotIS databases. Data and specimens were anonymised by using only their designated laboratory case reference. Ethical approval (ACP0000174) was gained from the Nottingham Health Science Biobank Access Committee. A cohort of 176 primary MM cases was used for iFRET analysis as tissue microarrays (TMAs). Supplementary Table 1 summarises the clinical parameters of the 176 patients. Tumours were fully surgically excised, formalin-fixed and paraffin embedded (FFPE) in tissue blocks. Tissue cores of 1mm diameter were selected by studying haematoxylin and eosin stained sections most recently cut from the FFPE tissue block. The location of cores to remove from the tissue block were selected by scanning the slides and using Pannoramic Viewer software (3DHisTech). Cores were removed from the FFPE tissue blocks using the TMA Grand Master (3DHisTech) and arrayed into new paraffin blocks.

### Antibodies and reagents

Monoclonal antibodies, mouse anti-PD-1 (ab52587), rabbit anti-PD-L1 (ab205921) and mouse anti-CTLA-4 (ab19792) were purchased from Abcam. Rabbit anti-CD80 (MBS2522916) was purchased from MyBioSource.

The Blockade Bioassay Kits for PD-1/PD-L1 (J1250) and CTLA-4/CD80 (CS186907) single cell assays and the anti-PD-1 blocking antibody (J1202) were purchased from Promega. Pierce endogenous peroxidase suppressor (35000), Signal Amplification kit (T20950) and Prolong diamond antifade mount (P36970) were obtained from Thermo Fisher Scientific. AffiniPure F(ab′)2 fragment donkey anti-mouse IgG and peroxidase-conjugated AffiniPure F(ab′)2 fragment donkey anti-rabbit IgG were purchased from Jackson Immuno Research Laboratories. ATTO 488 NHS ester was purchased and was conjugated to the AffiniPure F(ab′)2 IgG as described by Veeriah et al. 2014 [9]. Millicell® 8-well plates, (PEZGS0816) were purchased from Merck.

### Time-resolved amplified Förster resonance energy transfer (A-FRET) detected by fluorescence lifetime imaging microscopy (FLIM)

The conjugation of the chromophores to Fab fragments, which bind to the two primary antibodies, allowed the critical FRET distance of 10 nm or less to be maintained and provided the appropriate tool for measuring cell-cell interactions. Using a semi-automated, high throughput mfFLIM (FASTBASE Solutions S.L), a mapping file was created, which mapped each region of interest according to its position on the slide (Veeriah et al., [9]). Phase lifetimes, average intensities and lifetime images were calculated automatically and translated to an excel spreadsheet. A decrease of donor lifetime (*τD)* in the presence of the acceptor chromophore (*τDA*) is indicative of resonance energy transfer. FRET efficiency (E_f_ %) values were calculated using the following equation, where *τD* and *τDA* are the lifetimes of the donor in the absence and presence of the acceptor, respectively.

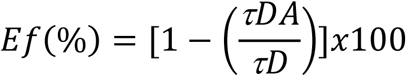

### Statistical analysis

Statistical analysis and Box and Whisker plots were performed using Origin Pro8. Statistical differences were calculated between groups using the Mann-Whitney U test (values indicated on the box and whisker plots). The Mann-Whitney U test is a nonparametric test, thus not assuming a normal distribution of results. Box and Whisker plots represent the 25–75% (box) and the 1–99 (whiskers) ranges. Statistical differences are indicated with p values ≤0.05. Kaplan-Meier survival analysis was performed using SPSS. Patients were ranked in order of their FRET Efficiency (interaction status) and split into the two groups; those with the highest 20% of FRET efficiencies, and those with the lowest 80%. The log-rank mantel cox test was carried out to determine significant differences between the groups.

### Immune-FRET (iFRET) assay for PD-1/PD-L1 interaction in cell culture

A PD-1/PD-L1 Promega Blockade Bioassay kit was identified as being adaptable to an amplified FRET protocol designed to measure PD-1/PD-L1 interaction. PD-L1 expressing CHO-K1 cells were seeded onto Millicell® 8-well plates and were incubated at 37 °C with 5% CO_2_ for 16 hours. The Promega J1201 anti-PD-1 blocking antibody was added to 4 wells at 25μg/ml final concentration to inhibit ligand-receptor interaction. PD-1 expressing Jurkat cells were subsequently seeded in all wells and the plates were incubated for 20 hours at 37 °C with 5% CO_2_. The unbound cells were removed and the plates washed three times for 5 minutes with phosphate buffered saline (PBS) before being fixed with 4% paraformaldehyde (PFA) for 12 mins. The PFA was then removed and the plates were washed three times for 5 min with PBS. All samples were incubated with endogenous peroxidase suppressor for 30 mins at room temperature (RT) before being washed with PBS. They were subsequently incubated with 1% (10mg/ml) Bovine Serum Albumin (BSA), for 1hr at RT before a further three PBS washes.

Primary antibody staining was carried out by adding mouse anti-PD-1, (1:100 in BSA), the donor only (D) readout condition. Meanwhile the donor acceptor (D/A) readout condition was labelled with both anti-PD-1 (1:100) and rabbit anti-PD-L1 (1:500). The plate was incubated overnight at 4°C before being washed twice with PBS containing 0.02% Tween 20 (PBST). Secondary Fab fragments were added, the D wells were labelled with anti-mouse FabATTO488 (1:100) and the D/A wells labelled with FabATTO488 (1:100) and anti-rabbit FabHRP (1:200). The plate was then incubated for 2h at RT before being washed twice with PBST and once with PBS.

Tyramide signal amplification (TSA) was performed on the D/A wells for 20 min in the dark, via the addition of Alexa594-conjugated tyramide diluted in amplification buffer (1:100) in the presence of 0.15% H_2_ O_2_. The D/A wells were washed twice with PBST and once with PBS to remove the tyramide. 5μl of Prolong Diamond anti-fade mount was added per well before being mounted with a coverslip.

### iFRET assay for CTLA-4/CD80 interaction in cell culture

The CTLA-4/CD80 Promega Blockade Bioassay kit was adapted to assess an Amplified-FRET protocol. CTLA-4 expressing Jurkat cells were first seeded onto Millicell® 8-well plate, before an anti-CTLA-4 antibody drug, ipilimumab (Qualasept), was added to 4 wells at 100μg/ml final concentration (100% inhibition). The CD80 expressing Raji cells were subsequently seeded and the cells were incubated for 20 h at 37 °C with 5% CO_2_. Unbound cells were removed by PBS washes. The cells were fixed, underwent endogenous peroxidase suppression and blocked with BSA as described previously in the PD-1/PD-L1 cell assay. The primary antibodies were added; the D wells were labelled with mouse monoclonal anti-CTLA-4 (1:100) and the D/A wells labelled with both anti-CTLA-4 (1:100) and rabbit polyclonal anti-CD80 (1:100). The rest of the protocol was conducted as described above for the PD-1/PD-L1 singe cell assay.

### iFRET assay for PD-1/PD-L1 interaction in formalin fixed paraffin embedded renal cell carcinoma (RCC) tissue

Human RCC tissue samples were provided by Cruces University Hospital, Bizkaia, Spain. Consecutive cross sections of tissues were mounted on separate slides to allow D and D/A antibody labelling. Samples were from 22 patients, from which 5 consecutive tissue section slides were provided. Of the 5 samples, 2 were available for D and 2 for D/A staining, while the remaining section was analysed using H&E staining to determine regions of immune infiltration. Immunohistochemistry with PD-L1 (SP-142, Ventana) was performed in Benchmark Ultra (Ventana) immunostainers following the specific protocol recommended by the customer.

For iFRET sample preparation, antigen retrieval was carried out using Envision Flex solution pH9 and a PT-Link instrument (Dako), where the slides were heated to 95°C for 20 minutes. Remaining paraffin was removed by PBS washes before containing tissue areas with a hydrophobic PAP pen border. 1-2 drops/slide of endogenous peroxidase suppressor were added and the slides were incubated in a humidified tray for 30 min at RT. The slides were then blocked with BSA and D slides labelled with anti-PD-1 while D/A slides were labelled with anti-PD-1 plus anti-PD-L1, following the previously described cell assay protocol.

### iFRET assay for PD-1/PD-L1 interaction in formalin fixed paraffin embedded malignant melanoma TMAs

Human malignant melanoma TMAs were provided by Nottingham University Hospitals, United Kingdom. Consecutive cross sections of tissues were mounted on separate slides to allow D and D/A antibody labelling. Samples from 176 patients, with two consecutive tissue section slides per patient were provided. Of the two samples, one was available for D and one for D/A staining. The primary antibodies used were anti-PD-1 and anti-PD-L1 following the same protocol as the renal tissue samples and the cell assay.

## Data Availability

Data will be available

## Acknowledgments

This work was supported in part by Department of Education, Basque Government-IT1270-19, Elkartek Grant (BG18) and the Spanish Ministry Grant (MINECO) PROJECTS of EXCELLENCE (BFU2015-65625-P). We would like to thank Pierre Leboucher for the automation of the multiple frequency domain FLIM. Prof Patel Poulam (Clinical Oncologist at Nottingham) and Prof Antoine Italiano for Clinical Discussions at the Bergonie Cancer Institute, Bordeaux, France.

## ABBREVIATIONS

PD-1: Programmed death receptor-1
PD-L1: Programmed death-ligand 1
CTLA-4: Cytotoxic T-lymphocyte-associated protein 4
CD80: Cluster of differentiation 80
FRET: Förster resonance energy transfer
FLIM: Fluorescence lifetime imaging microscopy
ccRCC: clear cell renal cell carcinoma
MSTS: multi-site tumour sampling
H&E: haematoxylin and eosin

